# Modelling of outbreaks of SARS-CoV-2 Omicron variants after easing Dynamic Zero-COVID strategy in mainland China

**DOI:** 10.1101/2022.12.22.22283841

**Authors:** Fujian Song, Max O. Bachmann

**Affiliations:** Norwich Medical School, University of East Anglia, Norwich, Norfolk, NR4 7TJ

**Author notes:** Correspondence to: Fujian Song.

## Abstract

**Background:** China lifted strict non-pharmacological interventions (NPIs) to prevent outbreaks of SARS-CoV-2 Omicron variants in December 2022. Relatively low levels of immunity and of vaccine booster coverage in the Chinese population raise concerns that future outbreaks will rapidly result in high rates of death and severe illness and will overwhelm health services.

**Methods:** This was a compartmental, discrete-time population dynamic model. It compared projected deaths and hospitalisation under various scenarios, including booster vaccine coverage and strictness of NPIs.

**Results:** We projected between 268,300 to 398,700 COVID-19 deaths, and peak numbers of hospitalised severe/critical cases between 3.2 to 6.4 per 10,000 population, before the outbreak wave recedes by February 2023. The COVID-19 deaths are reduced by 8% and 30%, respectively, under the weak and strict NPI scenarios, compared with the scenario without NPI measures. Early achievement of high coverage of three vaccine doses will further reduce COVID-19 deaths.

**Conclusion:** We projected fewer COVID-19 deaths and hospitalisations than some other models have. Rapid expansion of booster vaccine coverage will only be effective if combined with strict NPIs and/or the high coverage of booster vaccination could be achieved early.

## Introduction

Since the outbreak of the COVID-19 epidemic in China, strict non-pharmaceutical interventions (NPIs) have been implemented. The Dynamic zero-COVID strategy (DZCS), including mainly community lockdown and mass scale nucleic acid testing, has been successfully controlled sporadic outbreaks in China until November 2022. The reported total number of deaths from COVID-19 in China was only 5235 by 06/12/2022 (total population >1.4 billion), compared to 212,766 by 25/11/2022 in the UK (total population 68.75 million). However, by November 2022, it had become clear that the DZCS and its NPI measures are no longer able to control the transmission of SARS-CoV-2 Omicron variants, and that the strategy is unsustainable due to its social and economic burdens. Available evidence indicated that individuals infected with Omicron variants were mostly asymptomatic, with few severe/critical cases, particularly among those fully vaccinated.^1 2^ For these reasons, Chinese government started to adjust the COVID control strategy since 07/12/2022, to relax the stringent lockdown NPI measures, and to encourage older adults to have booster shots of COVID vaccines.^3^

Results of modelling studies have indicated that, without the strict NPI measures, the outbreak of SARS-CoV-2 Omicron variants could result in one million or more deaths in China.^4-6^ In this modelling study, we projected numbers of COVID-19 deaths and burdens to healthcare systems caused by the Omicron outbreak immediately after easing NPI measures in China.

## Methods

We revised and updated our model of effects of vaccination against COVID-19 in England,^7^ to represent future outcomes of Omicron outbreaks in China. It is a compartmental, discrete-time (day) population dynamic model (Supplementary Fig s1), implemented with computational language R.^8^ The population are classified into groups by age and COVID-19 vaccination status. The main infection compartments are susceptible, exposed, infectious, and recovered. The population of China is assumed to be susceptible to SARS-CoV-2 Omicron variant infection at the beginning of the simulation (01/11/2022), including individuals previously vaccinated against SARS-CoV-2. The exposed individuals are not infectious during the early incubation period, but start to be infectious before the onset of symptoms. Individuals infected may have no symptoms (asymptomatic), or palpable symptoms (symptomatic). Symptomatic patients are further classified into two categories: non-severe and severe/critical cases. Asymptomatic and non-severe cases may be isolated at home or not, and severe/critical symptomatic cases will need to be hospitalised, including those being admitted to intensive care units (ICUs). Individuals recovered from Omicron variant infection will develop immune responses against infection. In this short-term modelling study, the waning of protective immunity from natural infection and vaccination is not considered. It is assumed that previous vaccination against SARS-CoV-2 won’t protect vaccinated individuals from the infection by Omicron variant. However, vaccination will reduce infection severity and infection fatality.^2^

We estimated and calibrated the key infection transmission parameters based on a basic reproduction number R0=8.3, ^6^ and parameters used in a recent modelling study of Omicron outbreaks in China (Supplementary Table s1-s5).^4 6^ The primary outcomes are COVID-19 deaths and severe/critical cases who need hospitalisation following the COVID-19 Omicron infection. The age-specific IFR and ISR of unvaccinated individuals was estimated according to data on the SARS-CoV-2 Omicron outbreak in Shanghai,^1^ and effectiveness of vaccination with one, two and three doses of CoronaVac inactivated vaccine from Hong Kong.^2^ We used proportions of old people vaccinated with one, two and three doses of vaccines in China according to a paper by Zang et al^9^ and official information release.^10^

Although the Chinese government started to ease the Zero-COVID measures from 07/12/2022,^3^ it is unlikely that social activities could return to normal immediately, and the public are still very cautious going to public places and face masker wearing remains universal. In addition, the proportion of adults vaccinated with booster shot (three doses) will continue to increase in China. Because the rigorousness of NPI measures in coming weeks remains unclear, we compared several hypothetical scenarios with a range of different measures of NPI strictness after 07/12/2022 (Supplementary Table s5) and the booster vaccination coverage achieved by 31/01/2023 (Supplementary Table s2).

## Results

The projected COVID-19 deaths from 01/10/2022 to the end of the Omicron outbreak by February 2023 are from around 268,306 to 398,740 (Table 1 and Fig. 1a). The number of COVID-19 deaths ranges from 190 to 283 per million population. The daily peak number of hospitalised severe/critical cases ranged from 3.2 to 6.4 per 10,000 population (Table 1 and Fig. 1b). As expected, stricter NPI measures are associated with more delayed and flattened outbreak peaks, and the outbreaks are projected to end by February 2023 (Fig. 1). The COVID-19 deaths are reduced by 8-9% and 30%, respectively, under the weak and strict NPI scenarios, compared with the scenario without NPI measures. Compared with the moderate coverage of booster vaccination by 31/01/2023, the higher coverage reduces COVID-19 deaths by 4.6%, 4.3% and 3.6%, respectively, under the strict, weak and no NPI scenario. We also conducted an analysis by assuming an earlier reaching of high coverages of booster vaccination by 10/12/2022, rather than by 31/01/2023 as assumed in the above analyses. If so, the total COVID-19 deaths and the peak number of hospitalised cases could be further reduced by 6-8%.

**Table 1.** Projected total COVID-19 deaths, daily peak no. of severe cases hospitalised in China. NPI measures after 07/12/2022 from strict (NPI-2), weak (NPI-1), to none (NPI-0), and their corresponding initial reproduction numbers were 2.51, 3.06, and 4.00 respectively. Booster vaccination coverage by 31/01/2023 is assumed to be moderate or high (see Supplementary Table s2)

| Scenario | NPI strictness | Booster vaccine coverage by 31/01/2023 | Cumulative COVID-19 deaths | COVID-19 deaths per million population | Daily peak hospitalised COVID-19 patients | Peak hospitalised patients per 10,000 population |
| --- | --- | --- | --- | --- | --- | --- |
| NPI-21 | Strict | Moderate | 281,279 | 199.5 | 470,757 | 3.3 |
| NPI-11 | Weak | Moderate | 364,940 | 258.9 | 681,407 | 4.8 |
| NPI-01 | None | Moderate | 398,740 | 282.8 | 904,707 | 6.4 |
| NPI-22 | Strict | High | 268,306 | 190.3 | 454,037 | 3.2 |
| NPI-12 | Weak | High | 349,327 | 247.8 | 657,444 | 4.7 |
| NPI-02 | None | High | 384,308 | 272.6 | 875,852 | 6.2 |

**Fig. 1:**
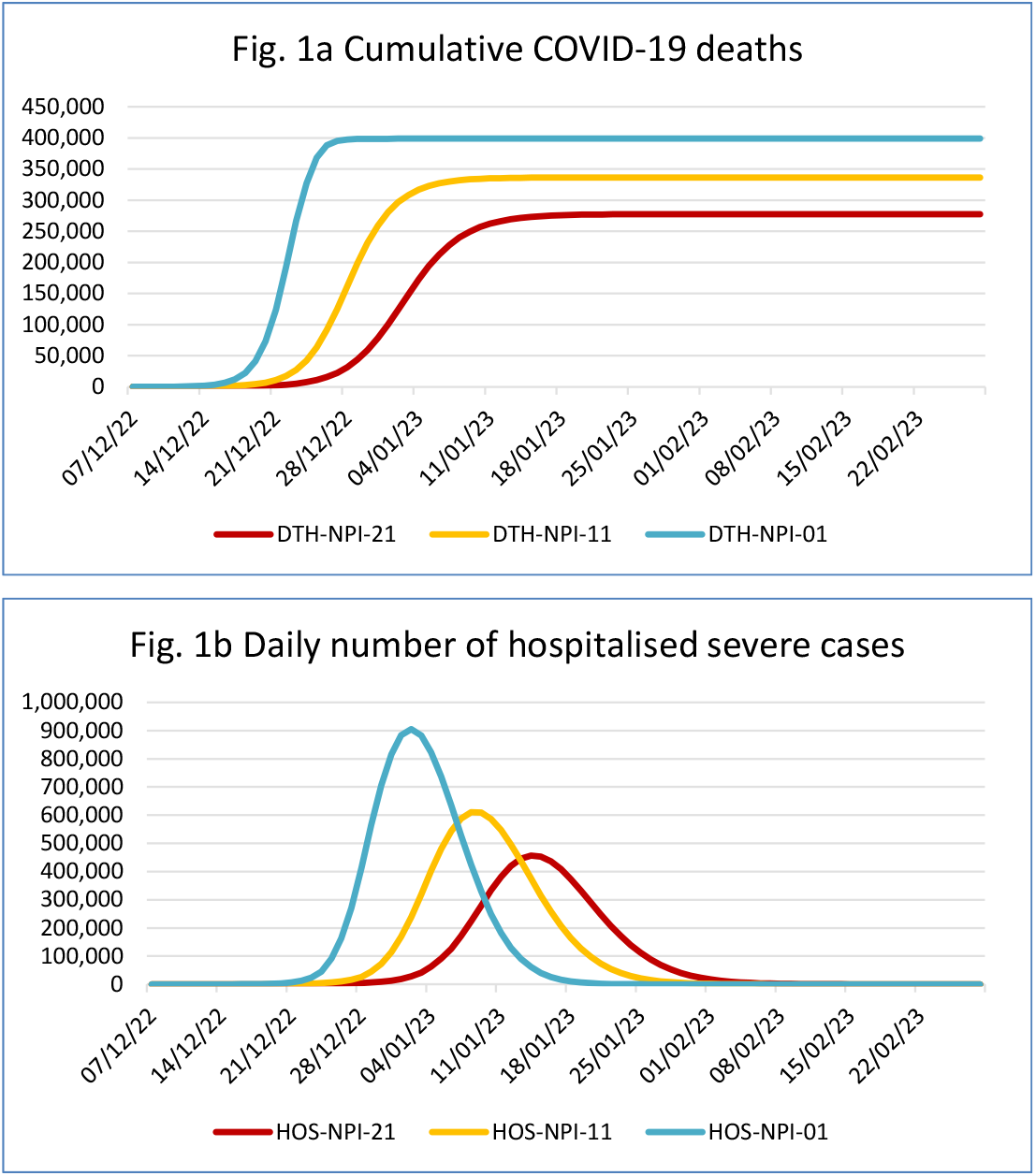
Projected cumulative COVID-19 deaths (Fig 1a) and daily number of hospitalised severe/critical cases (Fig 1b) under different strictness of non-pharmaceutical intervention measures after easing zero-COVID policy in China. NPI measures after 07/12/2022 from strict (NPI-2), weak (NPI-1), to none (NPI-0); assumed moderate coverage of booster vaccinations by 31/01/2023.

## Discussions

Our modelling study projected COVID-19 deaths after easing zero-COVID strategy in China, which range from 268,300 to 398,700, smaller than some other studies.^5 6 11^ We projected that the cumulative number of COVID-19 deaths by February 2023 are 190-283 per million population, compared to 568-770 per million by Leung et al,^5^ after lifting the zero-COVID measures. However, the total COVID-19 deaths in China was estimated to be from 225,453 to 293,127 by 01/04/2023 by IHME,^12^ which is smaller than ours. Differences in data and assumptions used may cause differences in the estimated COVID-19 deaths. In addition, the diagnostic criteria of the COVID-19 deaths may be different across countries and over time. COVID-19 deaths in our study is in line with the diagnostic criteria of COVID-19 deaths in Shanghai between February and June 2022, given the data source of infection fatality rates we used.^1^

The projected daily peaks of hospitalised severe/critical cases range from around 454,000 to 905,000, during the Omicron outbreak, which are below the number of hospital beds for respiratory illness (3.1 million) in China.^4^ A proportion of hospitalised COVID-19 patients will need ICU treatment, and the currently available ICU beds (n=64,000) may not be sufficient. It should be noted that, additional to the Omicron outbreak, the burden on hospital beds may further increase in the winter months due to influenza and other respiratory infections.

Efforts to increase booster vaccination in older adults will reduce harmful outcomes of the Omicron outbreak. Stricter NPI measures may acquire time for increasing the coverage of booster vaccination. Because of great transmissibility of SARS-CoV-2 Omicron variants (R0 around 8), the effective reproduction number after easing the zero-COVID policy is Rt=2.53 for the strictest NPI scenario used in this study. Therefore, the full benefit of incremental improvement in booster vaccination are difficult to achieve under the assumed NPI measures. If the higher coverage of booster vaccination could be achieved earlier by 10/01/2023 rather than by 31/01/2023, the COVID-19 deaths and hospitalisation burden could be further reduced by 6-8%.

China is a huge country with great diversities geographically and socioeconomically. Key estimates of infection fatality and severity risk were based on data from Shanghai, in which healthcare systems have better equipment and staff resources, compared with many other regions in China. In addition, new SARS-CoV-2 variants may emerge, which should be closely monitored. Therefore, results of this study may not be appropriate for specific regions or in future outbreaks. Outbreaks may occur early in major cities, and relatively later in remote areas. Optimising the booster vaccination of the population as early as possible, and maintaining appropriate NPI measures as strict as is feasible, will reduce fatalities and burdens on healthcare systems caused by the inevitable Omicron outbreaks.

## Data Availability

All data relevant to the study are included in the article or in supplementary information. Computational R code is available from FS.

## Contributors

FS designed, developed the model, retrieved data for estimating parameters, conducted computational calculations, and prepared the draft manuscript. MOB provided methodological support, helped interpret results and critically revised the draft manuscript. FS accepts full responsibility for the work and/or the conduct of the study, had access to the data, and controlled the decision to publish.

## Competing interests

No competing interests declared.

## Funding

There are no specific funding received for this study

## Data sharing statement

All data relevant to the study are included in the article or uploaded as online supplementary information. Computational R code is available from FS.

## Supplementary Files

### 1. Model structure and status

We revised and updated our existing model for vaccination against COVID-19 in England,^7^ using recently available data on Omicron variant of SARS-CoV-2 transmission and related health outcomes, to estimate the impact of changes in COVID-19 control policy in China.

It is a compartmental, discrete-time (day) population dynamic model (Supplementary Fig s1), implemented with computational language R.^8^ In this short-term modelling study, we assumed that the population will be constant, with balanced deaths and births, except deaths from COVID-19. Population in China are assumed to be susceptible to SARS-CoV-2 Omicron variant infection at the beginning of the simulation (01/10/2022), including individuals previously vaccinated against SARS-CoV-2. Susceptible individuals may be infected by contacting infectious individuals, and the infection status is changed from “susceptible” (SU) to “exposed” (EX). The exposed individuals are not infectious during the early incubation period, but start to be infectious before the onset of symptoms. Individuals infected with SARS-CoV-2 virus may have no symptoms (ASYM), and palpable symptoms (symptomatic or clinical infections). Asymptomatic individuals can spread SARS-CoV-2 virus before recovery, although the transmission risk may be lower than symptomatic patients. Symptomatic patients are further classified into two categories: non-severe and severe/critical symptomatic cases. Asymptomatic individual may be isolated at home (ASYMq) or not (ASYM0). Non-severe symptomatic cases may be isolated (SYMq) or not (SYM0), and severe/critical symptomatic cases will need to be hospitalised (HOS) (including those being admitted to intensive care units (ICU). Symptomatic patients are infectious and can transmit the virus to susceptible people before being isolated, hospitalised or recovered. We assume that hospitalised patients (HOS) are well isolated and no longer able to spread the virus to the susceptible population, although infectious patients who are self-isolated at home may transmit virus to household contacts.

Individuals may recover from infection of SARS-CoV-2 (RE). Individuals recovered from Omicron variant infection will develop immune responses against infection. In this short term (<6 months) modelling study, the waning of the protective immunity is not considered. It is assumed that vaccination against SARS-CoV-2 won’t protect vaccinated individuals from being infected with Omicron variant. However, infection severity and fatality risk will be reduced among vaccinated individuals.

**Fig s1:**
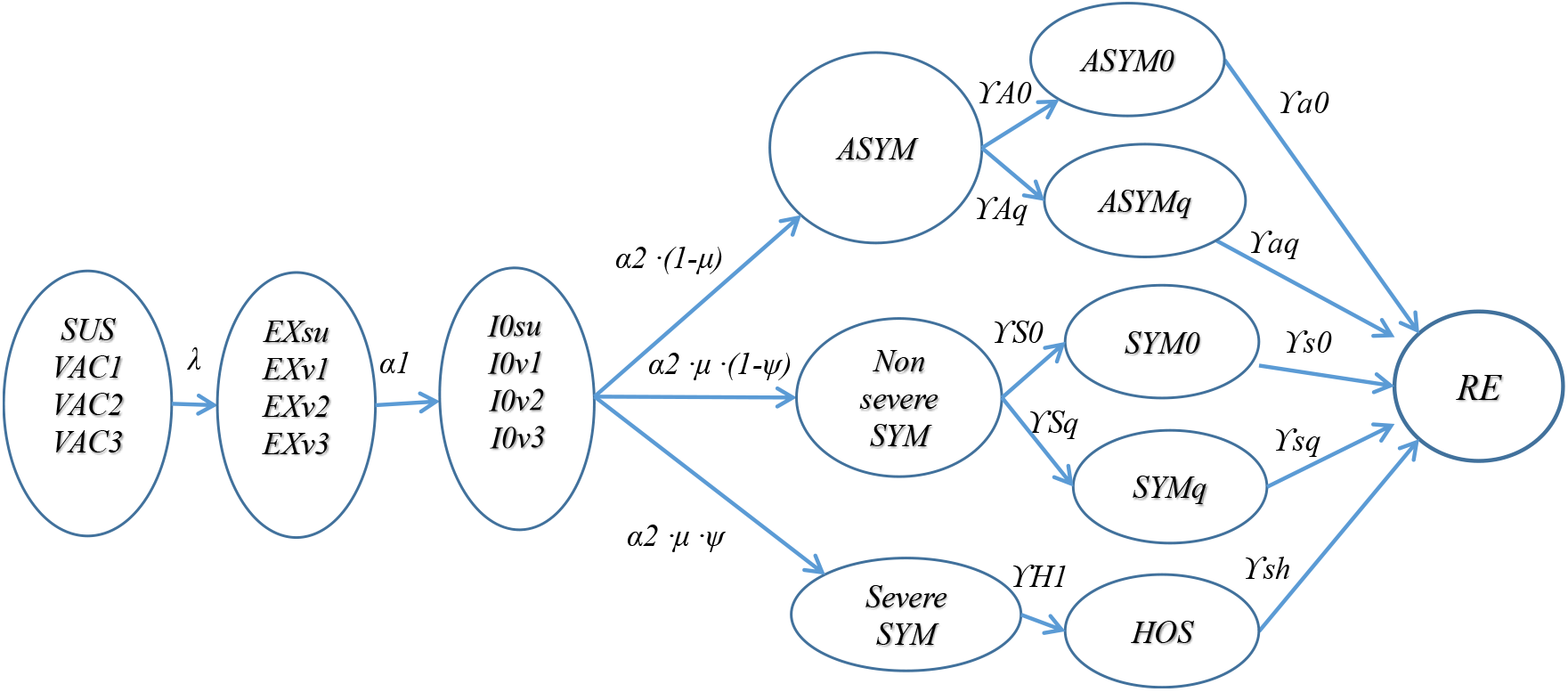
Modelling COVID-19 (omicron) epidemics in China – main compartments and transitions across status.

#### Definitions of compartmental variables in Fig s1

- SUS: susceptible individuals
- VAC1: vaccinated, dose-1
- VAC2: vaccinated, dose-2
- VAC3: vaccinated, dose-3
- EXsu: exposed susceptible individuals, not infectious
- Exv1: exposed vaccinated (dose-1), not infectious
- Exv2: exposed vaccinated (dose-2), not infectious
- Exv3: exposed vaccinated (dose-3), not infectious
- I0su: infectious, before symptom onset
- I0v1: infectious, before symptom onset, vaccinated dose-1
- I0v2: infectious, before symptom onset, vaccinated dose-2
- I0v3: infectious, before symptom onset, vaccinated dose-3
- ASYM: infectious, asymptomatic, with no or very mild symptoms
- ASYM0: asymptomatic cases not quarantined
- ASYMq: asymptomatic cases quarantined at home
- SYM: infectious, symptomatic cases
- SYM0: non-severe symptomatic patients who are not quarantined
- SYMq: non-severe symptomatic patients quarantined
- HOS: severe symptomatic patients who are hospitalised
- RE: recovered from covid-19 infection

#### Transition parameters in Fig s1

- λ: Force of infection (*λ*) measures the risk (probability) of infection transmission.
- α1: rate of progressing from being exposed to being infectious.
- α2: rate of progressing from being asymptomatic infectious to symptomatic.
- *μ*: proportion of infected individuals who will be symptomatic; age-specific, and vaccine dose-specific
- *?:* proportion of severe/critical cases among symptomatic individuals, age-specific, and vaccine dose-specific
- *pAq:* proportion of quarantined individuals among asymptomatic individuals
- *pSq:* proportion of quarantined individuals among non-severe symptomatic individuals
- γa0: rate of recovering for asymptomatic individuals
- γs0 rate of recovering for non-severe symptomatic cases
- γsq: rate of being isolated in symptomatic patients
- *γh1*: rate of being hospitalised for severe symptomatic cases
- *γsh*: rate of being hospitalised for severe symptomatic patients

### 2. Transition parameters and distribution of infectious period

In Fig s1, force of infection (*λ*) measures the risk of infection,^13^ which is a function of transmission rate (η) and the prevalence of existing infectious individuals (*I*) among the population (N): *λ=η·I/N*. The transmission rate *η* in the discrete-time model can be defined as the average number of new infected individuals generated daily by an infected person. That is, *η=Rt/T*, in which *Rt* is effective reproduction number and T is the average infectious period for infected individuals. We calculated *η* as a function of the number of daily contacts per person (c), and the risk of transmission per contact between a susceptible and an infected individual (*q*): η=c\**q*.^14 15^ For estimating age-specific transmission, *q* can be estimated according to known basic reproduction number (*R0*) and age-specific contract matrix, as R0 is the largest eigenvalue of the next generation matrix.^15^

The transition rate between model’s compartments in infectious models is often assumed to be constant, calculated by 1/x, in which “x” is the average period that subjects remain before the transition to the model’s next compartments.^13^ Therefore, the infectious period in standard SIR or SEIR models is usually assumed to be exponentially distributed, with some limitations of the use of exponentially distributed infectious period.^16 17^ In this study, we assumed that the transition probability between model’s compartments are based on gamma distributed periods that individuals remain in a compartment.^18^ The transition probability (y) at t is: *y*_*t*_ = (*cg*_*t*_ − *cg*_*t*−1_)/(1 − *cg*_*t*_), where *cg*_*t*_ is the gamma cumulative probability by the end of t. Given mean and shape (k) parameters, the gamma distribution based transition probability is used to estimate the number of individuals moving between two status.

### 3. Parameterisation and data sources

**Table s1:** Population, proportion of symptomatic, severe/critical cases, and infection fatality risk (unvaccinated) Data sources: Cai et al (2022),^4^ and Chen et al (2022).^1^

| Age | Population<sup>4</sup> | Symptomatic rate<sup>4</sup> | ISR(%)<sup>1</sup> | IFR(%)<sup>1</sup> |
| --- | --- | --- | --- | --- |
| 0-2 | 46730333 | 0.0716 | 0.0040 | 0.0010 |
| 3-11 | 155500009 | 0.0716 | 0.0040 | 0.0010 |
| 12-17 | 94764080 | 0.0716 | 0.0040 | 0.0010 |
| 18-24 | 104015331 | 0.0887 | 0.0400 | 0.0100 |
| 25-29 | 91847332 | 0.0887 | 0.0400 | 0.0100 |
| 30-34 | 124145190 | 0.0887 | 0.0400 | 0.0100 |
| 35-39 | 99012932 | 0.0887 | 0.0400 | 0.0100 |
| 40-44 | 92955330 | 0.1209 | 0.2800 | 0.0700 |
| 45-49 | 114224887 | 0.1209 | 0.2800 | 0.0700 |
| 50-54 | 121164296 | 0.1209 | 0.2800 | 0.0700 |
| 55-59 | 101400786 | 0.1209 | 0.2800 | 0.0700 |
| 60-64 | 73382938 | 0.1589 | 1.1400 | 0.3100 |
| 65-69 | 74005560 | 0.1589 | 1.1400 | 0.3100 |
| 70-79 | 80852004 | 0.1589 | 1.1400 | 0.3100 |
| 80+ | 35777716 | 0.1589 | 5.5500 | 2.2200 |

**Table s2:**
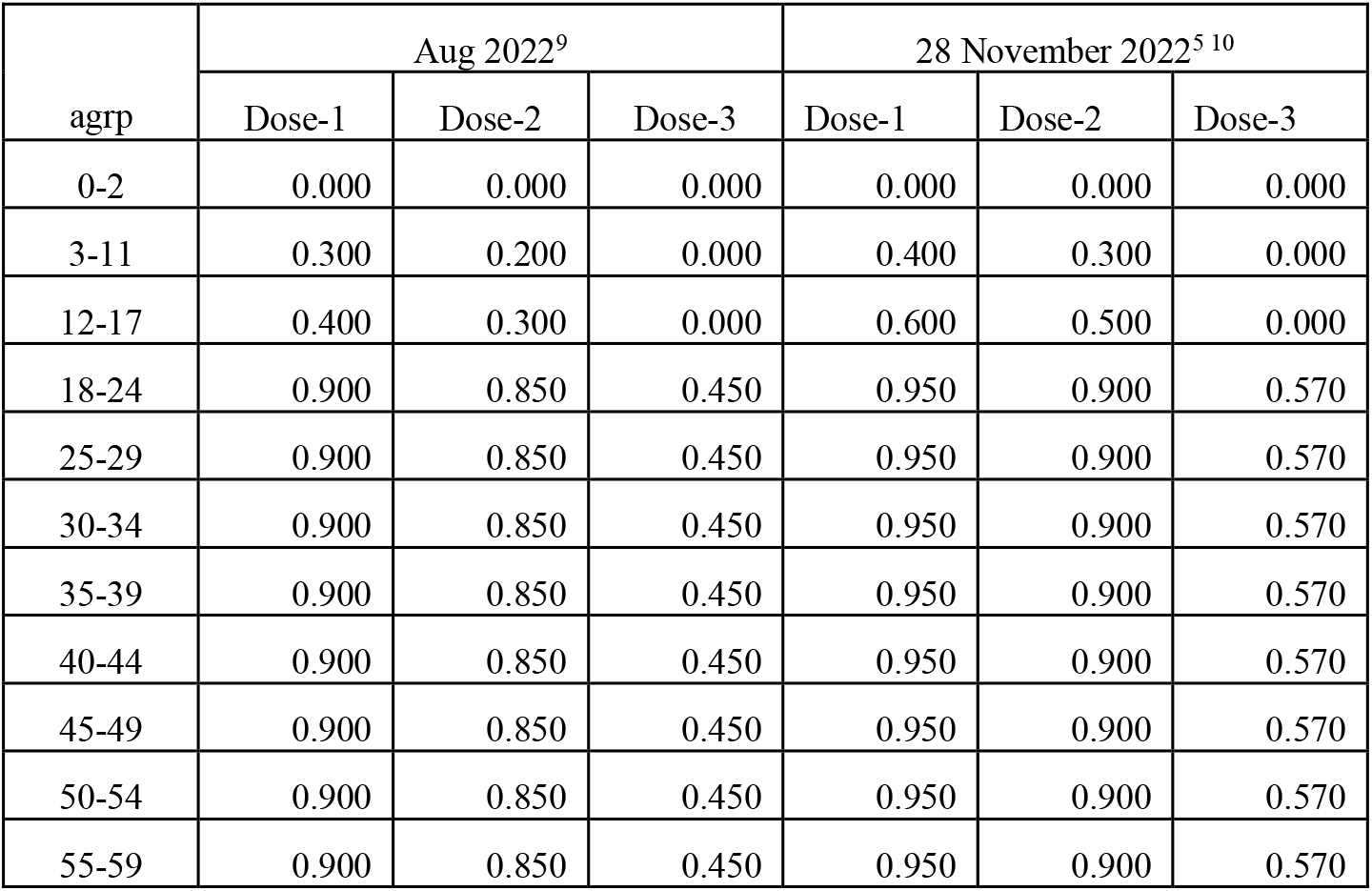
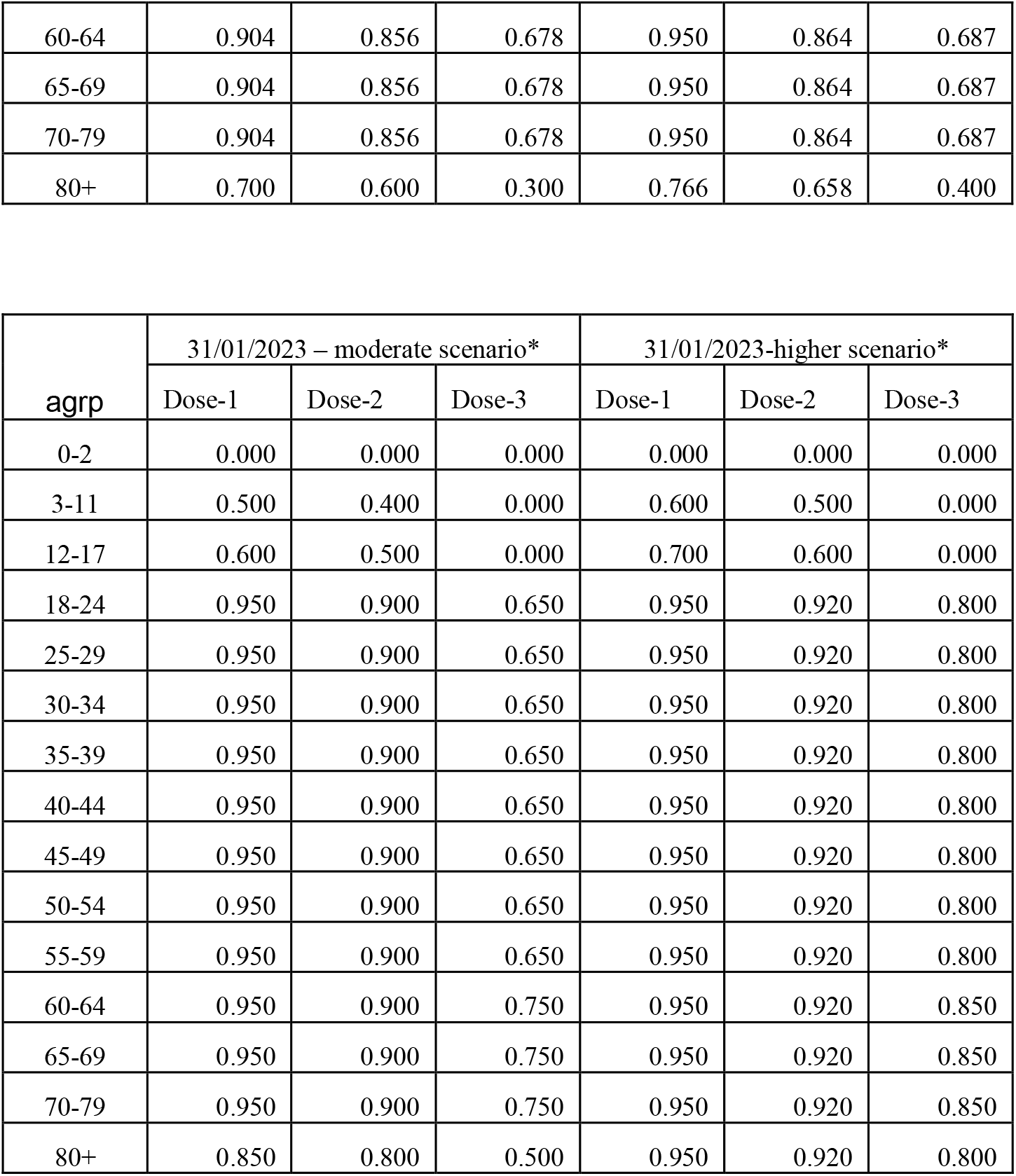
Proportions of individuals received one, two and three doses of SARS-CoV-2 vaccines. Data sources: Zang et al,^9^ State Council,^10^ and estimated*. The vaccination coverage is defined as the proportion of individuals vaccinated >14 days before.

| agrp | Aug 2022<sup>9</sup> |  |  | 28 November 2022<sup>5 10</sup> |  |  |
| --- | --- | --- | --- | --- | --- | --- |
|  | Dose-1 | Dose-2 | Dose-3 | Dose-1 | Dose-2 | Dose-3 |
| 0-2 | 0.000 | 0.000 | 0.000 | 0.000 | 0.000 | 0.000 |
| 3-11 | 0.300 | 0.200 | 0.000 | 0.400 | 0.300 | 0.000 |
| 12-17 | 0.400 | 0.300 | 0.000 | 0.600 | 0.500 | 0.000 |
| 18-24 | 0.900 | 0.850 | 0.450 | 0.950 | 0.900 | 0.570 |
| 25-29 | 0.900 | 0.850 | 0.450 | 0.950 | 0.900 | 0.570 |
| 30-34 | 0.900 | 0.850 | 0.450 | 0.950 | 0.900 | 0.570 |
| 35-39 | 0.900 | 0.850 | 0.450 | 0.950 | 0.900 | 0.570 |
| 40-44 | 0.900 | 0.850 | 0.450 | 0.950 | 0.900 | 0.570 |
| 45-49 | 0.900 | 0.850 | 0.450 | 0.950 | 0.900 | 0.570 |
| 50-54 | 0.900 | 0.850 | 0.450 | 0.950 | 0.900 | 0.570 |
| 55-59 | 0.900 | 0.850 | 0.450 | 0.950 | 0.900 | 0.570 |
| 60-64 | 0.904 | 0.856 | 0.678 | 0.950 | 0.864 | 0.687 |
| 65-69 | 0.904 | 0.856 | 0.678 | 0.950 | 0.864 | 0.687 |
| 70-79 | 0.904 | 0.856 | 0.678 | 0.950 | 0.864 | 0.687 |
| 80+ | 0.700 | 0.600 | 0.300 | 0.766 | 0.658 | 0.400 |

| agrp | 31/01/2023 – moderate scenario* |  |  | 31/01/2023-higher scenario* |  |  |
| --- | --- | --- | --- | --- | --- | --- |
|  | Dose-1 | Dose-2 | Dose-3 | Dose-1 | Dose-2 | Dose-3 |
| 0-2 | 0.000 | 0.000 | 0.000 | 0.000 | 0.000 | 0.000 |
| 3-11 | 0.500 | 0.400 | 0.000 | 0.600 | 0.500 | 0.000 |
| 12-17 | 0.600 | 0.500 | 0.000 | 0.700 | 0.600 | 0.000 |
| 18-24 | 0.950 | 0.900 | 0.650 | 0.950 | 0.920 | 0.800 |
| 25-29 | 0.950 | 0.900 | 0.650 | 0.950 | 0.920 | 0.800 |
| 30-34 | 0.950 | 0.900 | 0.650 | 0.950 | 0.920 | 0.800 |
| 35-39 | 0.950 | 0.900 | 0.650 | 0.950 | 0.920 | 0.800 |
| 40-44 | 0.950 | 0.900 | 0.650 | 0.950 | 0.920 | 0.800 |
| 45-49 | 0.950 | 0.900 | 0.650 | 0.950 | 0.920 | 0.800 |
| 50-54 | 0.950 | 0.900 | 0.650 | 0.950 | 0.920 | 0.800 |
| 55-59 | 0.950 | 0.900 | 0.650 | 0.950 | 0.920 | 0.800 |
| 60-64 | 0.950 | 0.900 | 0.750 | 0.950 | 0.920 | 0.850 |
| 65-69 | 0.950 | 0.900 | 0.750 | 0.950 | 0.920 | 0.850 |
| 70-79 | 0.950 | 0.900 | 0.750 | 0.950 | 0.920 | 0.850 |
| 80+ | 0.850 | 0.800 | 0.500 | 0.950 | 0.920 | 0.800 |

**Table s3:** Effectiveness of CoronaVac vaccination in severity and mortality reduction. Data source: McMenamin et al ^2^

| Age | Severity reduction |  |  | Mortality reduction |  |  | Mid/moderate disease |  |  |
| --- | --- | --- | --- | --- | --- | --- | --- | --- | --- |
|  | Dose-1 | Dose-2 | Dose-3 | Dose-1 | Dose-2 | Dose-3 | Dose-1 | Dose-2 | Dose-3 |
| <60 | 0.748 | 0.917 | 0.988 | 0.782 | 0.933 | 0.994 | 0.327 | 0.251 | 0.51 |
| 60_69 | 0.542 | 0.793 | 0.974 | 0.656 | 0.843 | 0.99 | 0 | 0 | 0.324 |
| 70_79 | 0.292 | 0.743 | 0.954 | 0.453 | 0.767 | 0.97 | 0 | 0 | 0.324 |
| 80+ | 0.39 | 0.582 | 0.973 | 0.448 | 0.63 | 0.979 | 0 | 0 | 0.324 |

**Table s4:** Estimates of transition parameters. Data sources: Song & Bachmann 2021,^7^ and Cai et al 2022.^4^

| Transition parameter | Value | Source |
| --- | --- | --- |
| Basic reproduction number | 8.3 | Based on the R0 (7.0 and 8.3) used in Leung et al.<sup>5</sup> |
| Serial interval (sd) | 3.0 (2.3) | Backer et al (2022)<sup>19</sup><br>For R package EpiEstim to estimate Rt. |
| Incubation period | 3.42 | Wu et al (2022)<sup>20</sup> |
| Incubation days: non-infectious | 2.42 (sd 1.556) |  |
| Incubation days: infectious | 1.00 (sd 1.00) |  |
| Average infectious days | 5 | Hakki et al (2022)<sup>21</sup> |
| Asymptomatic: Infectious days without isolation | 4.00 (2.00) |  |
| Asymptomatic: Infectious days before being isolated | 2.00 (1.414) |  |
| Symptomatic: Infectious days without quarantine | 4.00 (2.00) |  |
| Symptomatic: Infectious days before being quarantined | 1.00 (1.00) |  |
| Symptomatic: Quarantine to recovery days | 3.00 (1.732) |  |
| Severe/critical cases: days before hospitalisation | 2.20 (1.483) | Cai et al (2022)<sup>4</sup> |
| Length of hospital stay | 8.00 (2.828) | Cai et al (2022)<sup>4</sup> |
| Days from being infected to death | 11.40 (4.654) | Cai et al (2022)<sup>4</sup> |

### 4. Model verification/calibration

Based on the basic reproduction number (R0=8.3)^5^ and the pre-pandemic contact matrix in China,^4^ we estimated the risk of transmission per contact (*q=0*.*586*). We rum the basic simulation with 60 imported seeds on 01/10/2022, and calibrated parameters for NPI scaling factors in Table s5. The simulated incidence of new infected individuals was 26,600, which was close to the reported 25,321 on 06/12/2022, under the strict Dynamic Zero-COVID strategy in mainland China (http://www.nhc.gov.cn/xcs/yqtb/list_gzbd.shtml). Using R package EpiEstim and the simulated incidence of infected individuals, the effective reproduction number before 07/12/2022 was around Rt=1.41.

### 5. Projection scenarios

The Chinese government started to relax dynamic Zero-COVID policy from 07/12/2022.^3^ However, it is unlikely that the society and social activities could return to normal immediately, as some non-pharmaceutical intervention (NPI) measures will remain, and the public are still very cautious in going out. Face masker wearing is nearly universal, and access to some public places is still restricted.

Because the strictness of NPI measures in next many weeks is unclear, we assumed three NPI scenarios, respectively, with strict, weak and no NPI measures (Table s5). The early stage Rt corresponding to each of the assumed scenarios are, respectively, 2.53, 3.09, and 4.00.

Considering that the proportion of adults vaccinated with three doses will continue to increase, we assumed two scenarios with moderate or higher levels of booster vaccines in the population by 31 January 2023 (Table s2). Therefore, simulation scenarios are a combination of NPI measures after relaxing dynamic zero-COVID policy (Table s5) and whether the coverage of booster vaccine is moderate or higher by 31 January 2023 (Table s2).

**Table s5:** Non-pharmaceutical interventions measures (NPI) related parameters. Data sources: Contacts at home was assumed to be 20% of the general contacts. Risk of transmission per contact was reduced by q’ = q· (1-c), in which c is a coefficient, ranging from 0 to 1, as shown in the table.

| Parameters | Age | Non-pharmaceutical intervention measures |  |  |  |
| --- | --- | --- | --- | --- | --- |
|  |  | NPI -<br>zero COVID | NPI strict | NPI weak | No NPI |
|  |  | Before 07/12/2022 | from 07/12/2022 | from 07/12/2022 | from 07/12/2022 |
| Proportion of<br>quarantined<br>asymptomatic<br>cases | 0-17 | 0.920 | 0.600 | 0.200 | 0.050 |
|  | 18-59 | 0.920 | 0.600 | 0.200 | 0.050 |
|  | 60-69 | 0.920 | 0.600 | 0.200 | 0.050 |
|  | >70 | 0.920 | 0.600 | 0.200 | 0.050 |
| Proportion of<br>quarantined<br>symptomatic<br>cases | 0-17 | 0.980 | 0.800 | 0.600 | 0.200 |
|  | 18-59 | 0.980 | 0.800 | 0.600 | 0.200 |
|  | 60-69 | 0.980 | 0.800 | 0.600 | 0.200 |
|  | >70 | 0.980 | 0.800 | 0.600 | 0.200 |
| Scaling factor<br>for reduced $q$<br>and contacts | 0-17 | 0.910 | 0.700 | 0.500 | 0.00 |
|  | 18-59 | 0.890 | 0.600 | 0.400 | 0.00 |
|  | 60-69 | 0.930 | 0.700 | 0.500 | 0.00 |
|  | 70+ | 0.950 | 0.800 | 0.600 | 0.00 |

## Notes

### Competing Interest Statement

The authors have declared no competing interest.

### Funding Statement

This study did not receive any funding

